# LncRNA CDKN2B-AS1 C-Allele: A High-Risk Genetic Determinant in Cancer Susceptibility and Progression Impacting Immune Cell Modulation Dynamics with Therapeutic Implications

**DOI:** 10.64898/2026.01.04.25341444

**Authors:** Elisa Makdessi, Samar El-Hamoui, Fida El-Ayoubi, Malak Nabulsi, David Wehbe, Noha Ibrahim, Nadine Nasreddine, Nehman Makdissy

**Author notes:** **Corresponding author**: Prof. Nehman Makdissy; Lebanese University, Faculty of Sciences III, Ras-Maska, Lebanon. These authors contributed equally to this work (Samar El-Hamoui, Fida El-Ayoubi).

## Abstract

Examining LncRNA-driven cancer biology, our study represents the first comprehensive investigation of the LncRNA CDKN2B-AS1 across multiple cancer types, revealing its genetic basis for cancer susceptibility, and associations with thromboembolic risks and immunological dynamics. Stratification across hematological and non-hematological cancers revealed predominant prevalence rates of the CDKN2B-AS1 C-allele, notably higher in solid tumor malignancies, peaking at 86.8% in colorectal carcinoma (CRC). CRC-C-carriers exhibited worse outcomes, marked by reduced overall survival, increased venous-thromboembolism, elevated frequencies of thromboembolic-associated gene polymorphisms and BRAF mutations, and decreased MSI-H. Key tumor progression aspects in CRC-C-carriers included an inflammatory profile (reduced Th2 and increased Th1/Th17 cells), angiogenesis and endothelial activity (elevated CD14+, CD31+, CD144+, and VEGFR2+ cells), decreases in tumor suppressor-CD146^+^ cells (elevated tissue-adherent TA-EPCs lacking CD146), and marked elevation in tumor activity markers (Ki-67, CEA, CA 19-9, EGFR, VEGF-A, PD-1, and CTLA-4). Palbociclib significantly improved progression-free survival relative to baseline, primarily through suppression of tumor cell proliferation, with a more pronounced clinical and biomarker response observed in CDKN2B-AS1 non-C-allele carriers, whereas C-allele carriers exhibited an attenuated response consistent with relative treatment resistance. This underscores the multifaceted role of LncRNA CDKN2B-AS1, presenting it not just as a genetic determinant but also as a potential prognostic biomarker in cancer dynamics, paving the way for targeted personalized interventions, representing promising advancements in cancer therapeutics.

## 1. Introduction

Recent decades have witnessed an impactful shift in cancer biology, highlighting the pivotal involvement of long non-coding RNAs (lncRNAs) in tumor initiation, progression, and metastasis [1]. LncRNAs, a novel class of regulatory molecules, have been found to be dysregulated in a wide range of cancer types, often exhibiting aberrant expression patterns closely associated with clinical outcomes [2]. As their name suggests, lncRNAs do not directly translate into functional proteins; rather, they exert their influence by modulating the expression, stability, and localization of other genes through diverse mechanisms, including transcriptional regulation, post-transcriptional processing, and epigenetic modifications [3]. The complex nature of lncRNA-mediated gene regulation has positioned these non-coding RNAs as critical players in the complex networks that govern the hallmarks of cancer, such as uncontrolled cell proliferation, evasion of apoptosis, angiogenesis, and metastatic dissemination suggesting their potential as diagnostic, prognostic, and therapeutic biomarkers in oncology [4].

One genomic region that has garnered significant attention in the context of lncRNAs and cancer is the 9p21 locus [5]. This chromosomal region is known to harbor several tumor suppressor genes, including CDKN2A and CDKN2B, which play crucial roles in cell cycle regulation and tumor suppression [6]. Genetic variants and alterations within the 9p21 locus have been associated with a wide range of cancers, including leukemia, gliomas, ovarian, breast, and pancreatic cancers [7]. The lncRNA CDKN2B-AS1, also known as ANRIL, is one of the key players within the 9p21 locus. ANRIL plays a role in cancer development and progression [8], cell proliferation, apoptosis, inflammation, and extracellular matrix remodeling [9]. Elevated ANRIL levels are linked to lower survival rates and higher metastatic rates in CRC patients [10], and downregulation inhibits lymph angiogenesis and lymphatic metastasis [11], thus relating CDKN2B-AS1 to CRC progression [12] and being a risk locus [13]. Mechanistically, ANRIL functions as a molecular scaffold, interacting with poly-comb repressive complexes to modulate the expression of neighboring tumor suppressor genes, CDKN2A/B. CDKN2B encodes for the protein p15Ink4b that leads to cell cycle arrest by the inhibition of CDK4/6, thus impacting tumor cell proliferation. The C/G polymorphism within the ANRIL gene can contribute to the development and progression of various cancers. In fact, CDKN2B-AS1 carrying the G-allele promotes CDKN2B, while C-allele inhibits CDKN2B, leading to a converse effect of cell cycle progression [14,15]. Carriers of the C-allele have a higher risk of developing breast cancer and a lower survival rate in esophageal squamous cell carcinoma [16,17].

Primary complications in active or post-operative stages of cancer relate to the coagulation system, particularly is the thromboembolism. CDKN2B-AS1(C/G) polymorphisms’ association with increased risk of vascular diseases has been studied extensively [18–21], suggesting involvement in cancer-related thromboembolism [22]. It is crucial to target the hemostatic system by identifying thromboembolic risk biomarkers and as a target for anticancer therapy. Leading to activation of the coagulation system [23]and tumor progression [24], cancer cells activate vascular cells, release procoagulants, control fibrinolysis, and produce inflammatory and angiogenic cytokines, contributing to prothrombotic disorder of the vascular wall [25]. Cancer patients, especially those given systemic chemotherapy, are at much higher risk of thromboembolism due to abnormal blood clotting mechanisms [26].

In fact, despite the therapeutic progresses for effective anticancer therapy including chemotherapy, targeted therapies, immunotherapy, cell-based therapy, and sometimes radiation therapy, several treatments remain ineffective. Angiogenesis, inflammatory and highly proliferative tumor with certain genetic predisposition and genomic stability, might present challenges for certain types of treatments. Therefore, beyond the mutational signatures and molecular events that occur in cancers, the tumor microenvironment is central in understanding immunosurveillance and immunoediting in cancers [27], which describes interactions between the immune system and the tumor that allow cancer cells to evade immune surveillance, leading to tumor growth, chronic inflammation, and metastasis. The significance of how the inflammatory environment determines the immunological pathogenesis of cancer has been demonstrated by research on the activation patterns of immune cells and stromal cells [28]. Cancer prognosis is linked to immunological subsets, including central immune effector T cells (CD8^+^, CD25^+^), NK cells, and macrophages [28,29]. The production of Tregs and immunosuppressive factors like PD-1 [30], which puts CD8^+^ T-cells in a state of anergy [31,32], can hinder successful therapy, leading to the development of checkpoint blockade medications by preventing T-cell exhaustion or depleting Tregs [30]. Understanding the antitumor immune response and combining various strategies may enable effective immunotherapy for cancer treatment [30]. For instance, a chemo-immunotherapy trial stimulated human peripheral blood mononuclear cells (PBMCs) with colon cancer cells to produce cytotoxic T-cell lines specific for the disease [33]. Most therapies, including chemotherapy and biotherapy, have immunomodulatory effects that trigger immunogenic cell death and prevent immunosuppression.

Other than the genetic predisposition which may affect the immune system, circulating immune cells (CICs) and endothelial progenitor cells (EPCs) cells, and circulating infiltrated immune cells (CIICs), are promising for cancer therapy. CICs, such as CD4^+^ and CD8^+^ cells which are subsets of immune PBMCs provide insights into systemic immune responses and potential biomarkers for disease monitoring and treatment response. EPCs, subsets of bone marrow-derived stem cells implicated [34], in tumor angiogenesis and vascular remodeling, shed light on the tumor’s vascular network and metastatic potential, guiding anti-angiogenic therapies and prognostic evaluations. CIICs, comprising diverse immune cell subsets recruited to the tumor site, unveil mechanisms of immune evasion and inform immunotherapy strategies, aiding in patient stratification and treatment monitoring. Integrating assessments of these circulating entities and their relations with the LncRNA CDKN2B-AS1 variants, enables a complete understanding of cancer dynamics and facilitates the development of targeted interventions for improved patient outcomes.

While the dysregulation of CDKN2B-AS1 (ANRIL) has been reported in some cancer types, however CDKN2B-AS1 C/G prevalence across multiple types of cancer wasn’t studied previously. The majority of existing studies didn’t provide a holistic understanding of its significance in the context of diverse cancer biology. This knowledge gap limits our ability to establish ANRIL and its genetic variants as a robust prognostic and predictive biomarker, as well as to explore its potential as a therapeutic target in cancer management. At our knowledges, no previous study had reported in cancer the role of a potential LncRNA with CICs, EPCs, and CIICs, in one single study.

Therefore, the objective of this study is to investigate the LncRNA CDKN2B-AS1 C/G variant in cancer susceptibility, thromboembolic risk and immunological dynamics as predictive and prognostic biomarker. This study is to also provide a deeper understanding of lncRNA-driven cancer biology, evaluating the immune-inflammatory identity, the angiogenic factors and immune checkpoints, to guide personalized treatment strategies for cancer patients. Addressing this gap through a comprehensive investigation of CDKN2B-AS1 in diverse cancer contexts would significantly advance the field of cancer research and inform the development of more effective, personalized therapies, ultimately improving patient outcomes.

## 2. Material and Methods

### 2.1 Study design

The study involves a diverse group of 882 eligible outpatients aged 21-89 years, both males and females, diagnosed with various types of cancers, both hematological and non-hematological, as per the World Health Organization (WHO) cancer classification. Specifically, the study focuses on CRC, and the patients were diagnosed through colonoscopy and confirmed by evaluating the expression of specific genes and protein biomarkers. Aimed to cover a comprehensive understanding of the circulatory system and its crucial role in cancer, our investigations centered on the analysis of genes and proteins within the circulatory system relevant to thrombosis, thromboembolism, coagulation, etc. Concurrently, we conducted a thorough vascular assessment based on electrocardiography, echocardiography, and serum biomarker analysis to appraise vascular structure, function, and overall vascular health. The main assays utilized for screening and assessing biomarkers include immunohistochemistry, immunofluorescence microscopy, and PCR. Additionally, immunological markers were assessed using techniques such as flow cytometry, ELISA, and immune cell functional assays. Through the integration of these assessments, distinct patient subgroups were identified based on immunological, vascular, and cancer parameters. Selected CRC patients were further categorized into subgroups based on the cancer staging classified according to the AJCC TNM staging system, with specific exclusion criteria employed to ensure data integrity. These criteria ensure a focused examination of the targeted population and their background profiles concerning CRC, immunological factors, immunomodulation, vascular health, and related diseases. On the other hand, the staging of hematological cancers used the Ann Arbor staging system for Hodgkin lymphoma, the Lugano classification for non-Hodgkin lymphoma, and the French-American-British (FAB) classification for Leukemias. Patients have not received any form of therapy (radiotherapy, chemotherapy, immunotherapy, or cell-based therapy) at the time of diagnosis. The study outlines specific exclusion criteria to ensure the integrity of the data: inflammatory bowel diseases, cardiovascular diseases, coronary artery diseases, confirmed treated or untreated autoimmune, metabolic, diabetes, or neurological diseases. Additionally, individuals with evidence of cardiac, renal, bone, or cerebral damage, infections, myositis, familial polyposis, alcohol or smoking habits, colon-affecting food allergies, body mass index >30, significant weight loss within the last 2 years, history of abdominal surgeries, or pregnancy are also excluded.

The study was registered as a clinical study on clinicaltrials.gov with the identifier NCT06065592, titled “Exploring Cancer-Associated Thromboembolism Prognosis Biomarkers and Polymorphisms (CAT_PB)”, and conducted at the Haykel Hospital and the duration of the study was sixty months. The principal investigators enrolled patients and conducted follow-ups with all patients and collected comprehensive data on laboratory results, treatments, and outcomes from clinical records.

### 2.2 Serological and Blood Analyses

Blood samples were collected from both suspected subjects and selected patients, following standard procedures and laboratory operating protocols. A comprehensive list of assessments was performed on the blood samples, including measurements of complete blood count (CBC) or serology tests for albumin, globulin, total bilirubin, direct bilirubin, total proteins, creatinine, estimated glomerular filtration rate (eGFR), urea, uric acid, sodium, potassium, chloride, calcium, vitamin B12, vitamin D, blood sugar, hemoglobin A1c (HbA1c), total cholesterol (T-Chol), high-density lipoprotein (HDL), low-density lipoprotein (LDL), triglycerides (TG), prothrombin time (PT), activated partial thromboplastin time (aPTT), D-dimer, fibrinogen, troponin, antithrombin III (ATIII), protein C, protein S, serum complement C3 and C4, double-stranded DNA (dsDNA), antinuclear antibodies (ANA) and ANA profile, antineutrophil cytoplasmic antibodies (ANCA) and ANCA profile, lactate dehydrogenase (LDH), C-reactive protein (CRP), interleukin-6 (IL-6), iron, ferritin, homocysteine, creatine phosphokinase (CPK), complement C3, complement C4, antithrombin III (ATIII), lupus anticoagulant, antiphospholipid antibodies, anti-cardiolipin antibodies, alanine aminotransferase (ALT or SGPT), aspartate aminotransferase (AST or SGOT), alkaline phosphatase (ALP), gamma-glutamyl transferase (GGT), lipase, amylase, thyroid-stimulating hormone (TSH), hemoglobin, hematocrit, red blood cells (RBC), white blood cells (WBC), platelets, neutrophils, lymphocytes, monocytes, eosinophils, basophils, and erythrocyte sedimentation rate (ESR). For PCR assays, whole blood samples collected in EDTA tubes were utilized.

### 2.3 Human PBMCs Preparation

Twenty ml of fresh peripheral venous blood were collected into EDTA tubes. The blood was then diluted 1:1 with phosphate-buffered saline (PBS) and carefully layered over Ficoll-Paque PLUS, following which centrifugation (30 min, 400xg) at room temperature. The hPBMC layer was pooled and transferred into a 15-ml falcon tube. The sample was further diluted 1:1 with PBS and centrifuged again (5 min, 250xg) at room temperature. The resulting pellet was resuspended with 5 ml of PBS and subjected to centrifugation at 250xg for 5 min. The cell pellet was finally resuspended in 2 ml of PBS. Manual and flow cytometry-based cell counting methods were employed.

### 2.4 CDKN2B-AS1 Genotyping

For genotyping of CDKN2B-AS1, all individuals were genotyped for the chr9p21.3 rs1333049 polymorphism (C/G, Transversion Substitution) using the TaqMan SNP allelic discrimination genotyping assay (Applied Biosystems, Thermo Fisher Scientific, CA, USA; assay identification C__1754666_10, Cat# 4351379). A total of 20 ng of purified DNA were amplified in a final volume of 10 μL per well in 384 well plates with a PCR cycling protocol of initial denaturation at 95°C for 10 min, followed by 40 cycles at 92°C for 15 sec and 60°C for 1 min. Post PCR allelic discrimination was carried out by measuring allele-specific fluorescence on an ABI prism® 7500 Sequence Detection System (Applied Biosystems) using the ABI Prism SDS software version 2.1 (ABI). Genotyping was successful in 97% of rs1333049 polymorphism. TaqMan SNP Genotyping Assays required only three reaction components for PCR: purified genomic DNA, the assay solution, and TaqMan Genotyping Master Mix. Internal probes were: VIC™ (5’), FAM (5’), MGB (Minor Groove Binder) (3’), where labels (dye) were VIC (probe 1, to detect the allele C) and FAM (probe 2, to detect the allele G); The well contains a known template to generate a specific genotype call for an assay, specifying positive controls for VIC/VIC, VIC/FAM, or FAM/FAM. FAM™ and VIC® dye fluorescent values are normalized by the ROX passive reference dye. The context sequence [VIC/FAM] is: CATACTAACCATATGATCAACAGTT[C/G]AAAAGCAGCCACTCGCAGAGGTAAG.

For genotyping of thrombophilic-Related Gene Polymorphisms (TRGP, genes and polymorphisms associated with blood clot formation and thrombotic events, such as deep vein thrombosis, pulmonary embolism, and arterial thrombosis), and other Vascular-Related Gene Polymorphisms (VRGP, genes and polymorphisms involved in lipid metabolism, and inflammation), StripAssay assessment by multiplex PCR and reverse-hybridization were applied as previously described [35], utilizing Viennalab Biotechnology and SNP Biotechnology.

### 2.5. Biomarkers Analysis

Tumor biopsies were immunohistochemically analyzed using conjugated antibodies against CEA, CA 19-9, EGFR, VEGF-A, PD-1, CTLA-4, and Ki-67. KRAS and BRAF mutation analyses were performed on formalin-fixed, paraffin-embedded (FFPE) tissues following genomic DNA extraction (QIAamp DNA FFPE Tissue Kit, Qiagen), quantification (NanoDrop ND-1000), and PCR-based reverse-hybridization assays (KRAS XL and BRAF 600/601 StripAssay; ViennaLab). KRAS mutations were assessed across exons 2–4 (codons 12, 13, 59, 61, 117, and 146), and BRAF mutations at codons 600 and 601. Microsatellite status was determined by immunohistochemical evaluation of mismatch repair proteins (MLH1, MSH2, MSH6, and PMS2), with MSI defined by loss of nuclear expression of one or more MMR proteins in tumor cells.

### 2.7. Immunohistochemical Profiling of Tumor Markers and Immune Cell Infiltration

IHC was utilized to identify tumor markers, tumor-infiltrating immune cells (TIICs), and other pertinent markers in formalin-fixed paraffin-embedded tissue samples. Expert pathologists reviewed hematoxylin eosin-stained slides to confirm diagnoses. Automated immunostainers were employed to ensure consistency during the staining process, incorporating appropriate positive and negative controls. Markers investigated include, but are not limited to: immune cell markers such as CD3, CD4, CD8, CD5DIN, CD7, CD15, CD20, CD23, CD30, CD34, CD38, CD43, CD45, CD79a, and CD138; proliferation markers KI67 and Cyclin D1; oncogenes/tumor suppressor genes p53, b-2 microglobulin, bcl 2, bcl 6, Myeloperoxidase, Muramidase, PDL1, and HER2; hormone receptors Estrogen and Progesterone; and other pertinent markers TPS-A, CD10, CD17, E-cadherin, GATA3, and PAX5.

### 2.8. Immunophenotyping of CICs, EPCs, immune checkpoints, Th cells, and cytokines production

Flow cytometry was used to assess the immunophenotyping of cell surface markers expressed by CICs and EPCs, as well as viability and apoptotic index using 7AAD/Annexin V/PI staining, and immune checkpoint expression targeting CD152 (CTLA-4) and CD279 (PD-1). The analyses were performed as described previously [36,37]. Immunophenotyping was carried out using human conjugated Abs (Vioblue, FITC, APC, PerCP, PE, etc.) and isotype control Abs. 1×10^5^ cells were stained and analyzed by flow cytometry. Isotype and automated compensation were adjusted to minimize false positive fluorescence and spectral overlap of fluorochromes, respectively. Data based on 20,000 events were acquired from each sample in a user-defined gate and analyzed.

For Th1, Th2, and Th17 immunophenotyping assays, cells were fixed, permeabilized and stained using a mixture of CD4 PerCP-CY5.5, IFN-γ-FITC, IL-4-APC, and IL-17A-PE Abs according to the manufacturer’s instructions (Human Th1/Th2/Th17 Phenotyping kit, BD Biosciences). Analyses were realized by flow cytometry. The cytokines levels were assessed using the MACSPlex cytokine 12 kit (Miltenyi) following the manufacturer instructions.

For EPCs immunophenotyping, we utilized a sequential gating strategy to distinguish three distinct phenotypes. Initially, we gated on the CD45 marker to select the CD45-negative population, primarily composed of non-hematopoietic cells. Then, within this CD45-negative population, we further gated on the CD34 marker to isolate CD34-positive cells. Following this, we gated on the KDR (VEGFR-2) positive population, representing cells with endothelial lineage potential. Within the KDR-positive population, we then gated on the CD31 (PECAM-1)-positive cells, refining our selection to endothelial cells. Further gating based on CD144 and CD146 enabled the identification of three distinct phenotypes: Phenotype “A” (CD45^-^/CD34^+^/KDR^+^/CD31^+^/CXCR4^+^/CD144^-/^CD146^-^), Phenotype “B” (CD45^-^/CD34^+^/KDR^+^/CD31^+^/CXCR4^+^/CD144^+^CD146^-^), and Phenotype “C” (CD45^-^/CD34^+^/KDR^+^/CD31^+^/CXCR4^+^/CD144^+^/CD146^+^).

### 2.9. Treatments

Patients received chemotherapy and targeted therapy. Chemotherapy consisted of CAPOX administered in 3-week cycles for a total of 8 cycles (oxaliplatin 130 mg/m² intravenously on day 1, combined with capecitabine 1,000 mg/m² orally twice daily for 14 days followed by a 7-day rest). Targeted therapy consisted of Palbociclib (PAL) administered orally at 125 mg once daily for 21 consecutive days followed by 7 days off. Patients received either CAPOX alone or CAPOX combined with PAL (CAPOX as first-line treatment, with PAL administered as second-line therapy).

### 2.10. Clinical Response Evaluations in Treatments

Tumor regression after 2 cycles of treatment was assessed using CT scans for evaluation of either progressive or stable disease. The initial assessment included medical history, clinical examination, full laboratory analysis, and metastatic evaluation using chest CT, enhanced CT or MRI of the abdomen and pelvis, colonoscopy, and tumor biopsy. The clinical responses were assessed following the Response Evaluation Criteria in Solid Tumors version 1.1 guideline (RECIST version 1.1). Adverse events were graded according to the National Cancer Institute Common Terminology Criteria for Adverse Events (CTCAE), version 4.02 (CTCAE version 4.02).

### 2.11. Statistical analysis

Kaplan-Meier analysis was utilized to assess survival probabilities, and alongside this, Log-rank (Mantel-Cox) test, Logrank test for trend, Gehan-Breslow-Wilcoxon test, Median survival, Hazard Ratio (Mantel-Haenszel), and Hazard Ratio (Logrank) were conducted to comprehensively evaluate outcomes differences and trends in outcomes. For a parameter to be reported, at least four tests must be significant, ensuring robustness in the findings regarding the association between the studied parameter and outcomes in our study population. Additionally, Cox proportional hazards regression analysis was employed to assess the association between predictor variables and time-to-event outcomes, particularly focusing on the risk factors associated with mortality. Consistently, a significance level of α = 0.05 and two-tailed P values ensured vigorous statistical evaluation, contributing to an in-depth understanding of LncRNA CDKN2B-AS1 variants’ impact on patients.

## 3. Results

### 3.1. Genetic Analysis of LncRNA CDKN2B-AS1 Variants in Cancer Susceptibility: Highlighting High Prevalence of Risk Allele “C” in Solid Cancers

We screened 1,743 unrelated outpatients, of which 882 met the study criteria, exhibiting a male-to-female ratio (M/F) of 0.66 and a mean age of 49.5 ± 0.74 years, clinically suspected of having cancers (Table IA). Among them, 380 were diagnosed as cancer-free (CF group), while 502 were classified as cancer-positive (CP group).

**Table I:** Baseline Parameters of the Cohort and Prevalence of the CDKN2B-AS1 ‘C’ Allele in Diverse Cancer Types.

| **A - Baseline characteristics** | **Total** | **Cancer Free <sup>(1)</sup>** |  | **Cancer Positive <sup>(2)</sup>** |  |  |  |
| --- | --- | --- | --- | --- | --- | --- | --- |
| Total Population – *n* | 882 | 380 |  | 502 |  |  |  |
| Age, years (Mean ± SEM) | 49.5 ± 0.74 | 46.8 ± 0.94 |  | 54.7 ± 0.82 |  |  |  |
| Gender (male-to-female ratio (M/F)) | (0.66) | (0.47) |  | (0.81) |  |  |  |
| Race, white (%) | (100%) | (100%) |  | (100%) |  |  |  |
| **B - Occurrence of CDKN2B-AS1 “C” allele <sup>(3)</sup>** | **Total – *n*** | **non-C-carriers** |  | **C-carriers** |  | **OR** | **P** |
| **Total Population – *n* (%)** | 882 | 401 | (45.5) | 481 | (54.5) |  |  |
| Cancer-Free | 380 | 281 | (73.9) | 99 | (26.1) |  |  |
| Cancer-Positive | 502 | 120 | (23.9) | 382 | (76.1) | 9.04 | <.0001 |
| **Hematological Cancer <sup>(4)</sup>** | 56 | 27 | (48.2) | 29 | (51.8) | 3.05 | <.0005 |
| Lymphocytic Cancer | 33 | 13 | (39.4) | 20 | (60.6) | 4.37 | <.0001 |
| Myeloid Cancer | 23 | 14 | (60.9) | 9 | (39.1) | 1.82 | NS |
| **Non-Hematological Cancer** | 446 | 93 | (20.9) | 353 | (79.1) | 10.8 | <.0001 |
| **Gastrointestinal (GI) Cancers** |  |  |  |  |  |  |  |
| Lower GI - Colorectal Carcinomas <sup>(5)</sup> | 219 | 29 | (13.2) | 190 | (86.8) | 18.6 | <.0001 |
| Middle GI - Hepatobiliary Carcinomas <sup>(6)</sup> | 23 | 5 | (21.7) | 18 | (78.3) | 10.3 | <.0001 |
| Upper GI - Gastric, Pancreatic Carcinoma <sup>(7)</sup> | 16 | 7 | (43.8) | 9 | (56.2) | 3.65 | <.05 |
| **Genitourinary Cancers** |  |  |  |  |  |  |  |
| Ovarian & Other Gynecologic Carcinomas <sup>(8)</sup> | 49 | 10 | (20.4) | 39 | (79.6) | 11.1 | <.0001 |
| Prostate Carcinoma | 27 | 9 | (33.3) | 18 | (66.7) | 5.68 | <.0001 |
| Bladder Carcinoma | 23 | 10 | (43.5) | 13 | (56.5) | 3.69 | <.005 |
| **Other Solid Tumor Carcinomas** |  |  |  |  |  |  |  |
| Lung Carcinomas <sup>(9)</sup> | 35 | 7 | (20.0) | 28 | (80.0) | 11.4 | <.0001 |
| Breast Carcinoma | 51 | 15 | (29.4) | 36 | (70.6) | 6.81 | <.0001 |
| Brain Carcinoma (including Glioblastoma) | 3 | 1 | (33.3) | 2 | (66.7) | 5.68 | NS |

Initially, LncRNA CDKN2B-AS1 genotyping for the alleles (C) and (G) was conducted in CF and CP groups across 23 types of cancers (Table IB). CP patients exhibited a notably high frequency of the C-allele (0.76, denoted as C-carriers) and a lower frequency of the G-allele (0.24, denoted as non-C-carriers). The expression of the C-allele was significantly predominant in CP compared to CF (OR: 9.04, P<.0001).

Subsequent stratification into hematological and non-hematological cancers revealed distinct prevalence rates, highlighting the potential role of the C-allele in cancer susceptibility, particularly in solid tumors (0.79; OR: 10.8, P<.0001) compared to hematological malignancies (0.52; OR: 3.05, P<.0005). Notably, in hematological cancers, the C-allele exhibited significantly higher occurrence in lymphocytic malignancies (0.61; OR: 4.37, P<.0001), while in non-hematological cancers, its highest prevalence was observed in gastrointestinal carcinomas, notably colorectal carcinoma (CRC) (0.87; OR: 18.6, P<.0001), followed by lung cancers (0.80; OR: 11.4, P<.0001), and ovarian cancers (0.79; OR: 11.1, P<.0001).

Given its prevalent presence, especially in solid tumors, the C allele was designated the “Risk allele” for its association with cancer susceptibility across various types of cancers.

### 3.2. CRC Susceptibility, Mortality, and Thromboembolic Risk: Stratification by C-carrier Status

To explore the impact of the risk allele on CRC susceptibility, a cohort of 448 subjects was examined. Among them, 229 individuals formed the control group, while 219 subjects constituted the CRC group. Within the CRC group, patients were further classified based on the AJCC TNM staging system, covering stages 0-IV, representing the early (Stages 0 and I, CRC-ES-group, characterized by localized tumors), mid (Stage II, CRC-MS-group, indicating deeper infiltration) and advanced stages of CRC (Stages III and IV, CRC-AS-group, denoting distant CRC). Notably, all participants were non-hospitalized at the time of diagnosis and had not undergone any prior lines of therapy.

Data from the CRC group revealed significant insights (Table IIA): (1) the primary tumor was predominantly situated on the left side of the colon (59.8%), although 32.9% of patients exhibited right-side adenocarcinomas; (2) surgical interventions predominantly resulted in complete resection (60.3%), with 26.0% of patients undergoing partial resection; (3) the majority of CRC patients demonstrated a predominant tumor grade of moderately differentiated histology (79.0%), while a minority presented with poorly differentiated (12.8%) or well-differentiated (8.2%) histology; (4) over the 60-month study duration, aside from hospitalization related to therapy, 14.6% of patients required hospitalization, with an average length of stay (LOS) of 9.1 days, and only 1.8% were admitted to the intensive care unit (ICU); (5) the incidence of mortality within the CRC group was 10.9%.

**Table II:**
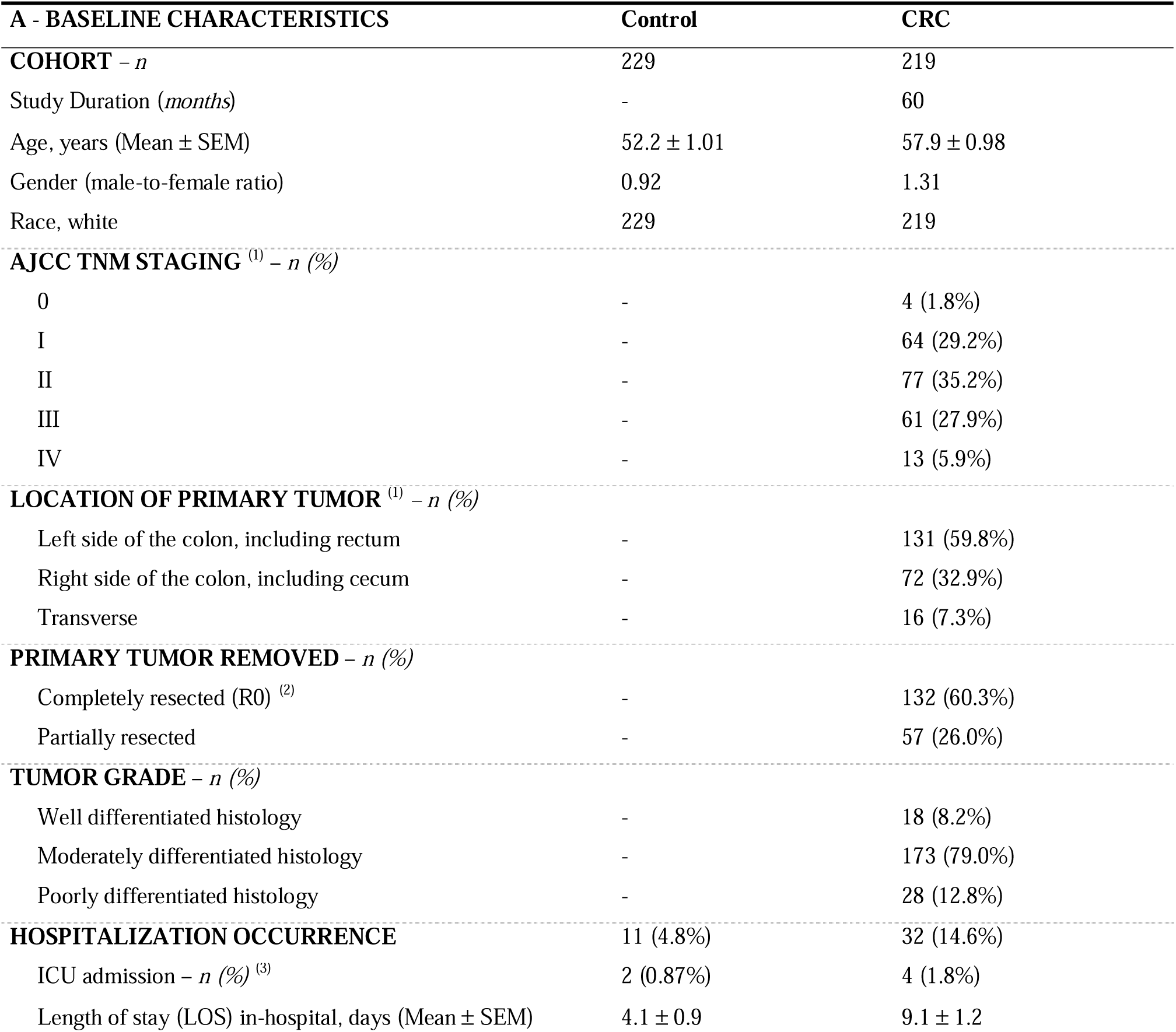
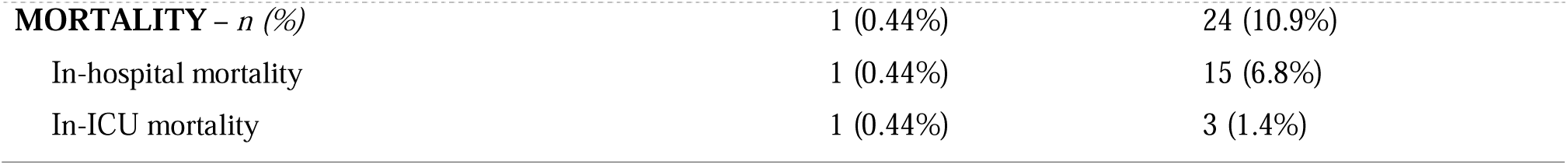
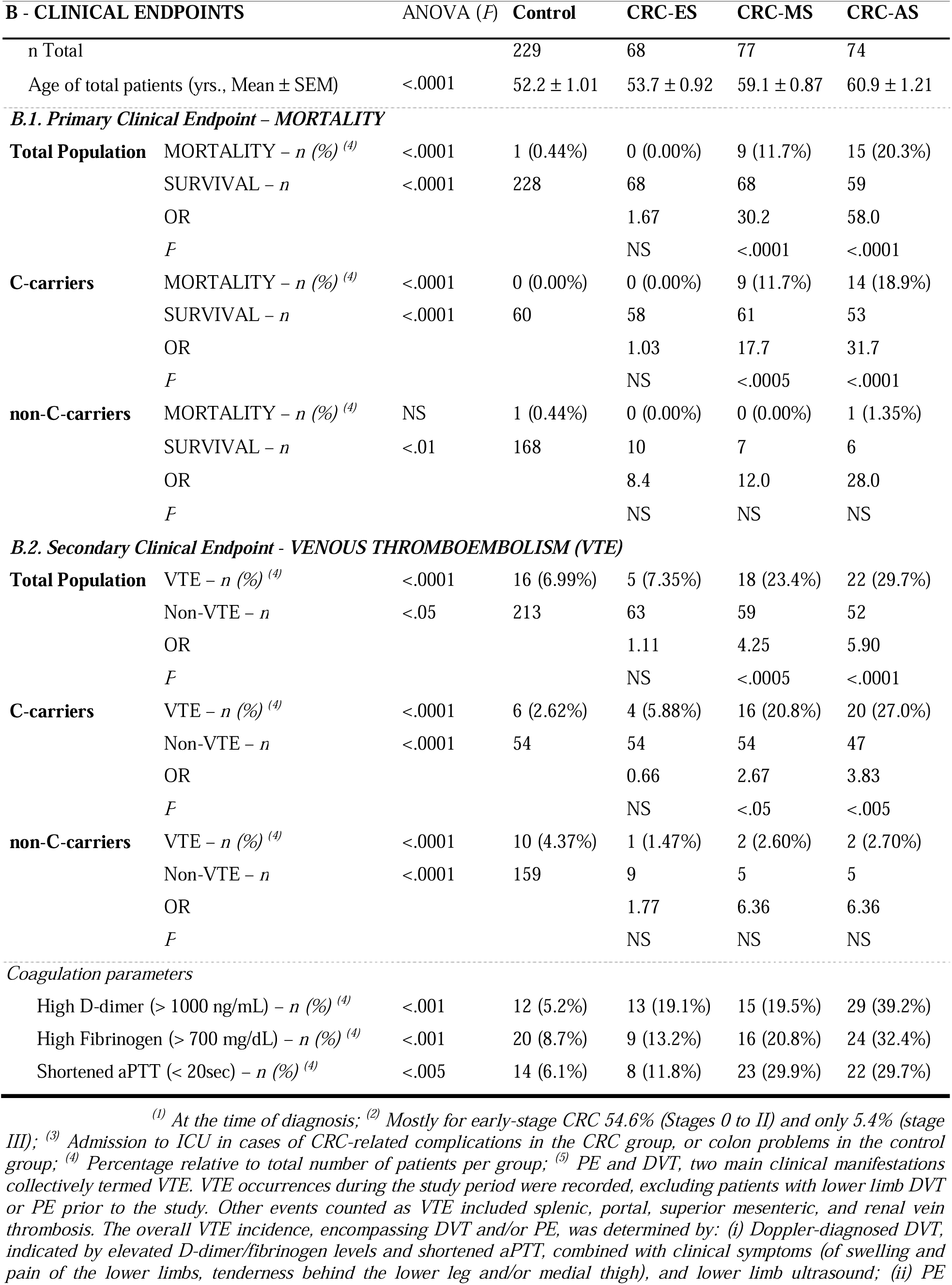
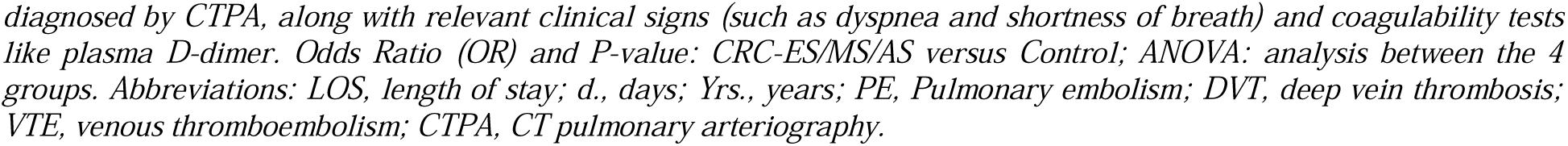
Baseline Characteristics and Clinical Endpoints of the Colorectal Cancer Cohort.

Significant differences were observed among the groups for the primary clinical endpoint of mortality (Table IIB1). Mortality rates were notably elevated in the CRC-MS (11.7%) and CRC-AS (20.3%) groups compared to the control (0.44%), indicating an increased risk of mortality with advanced staging of CRC. Among C-carriers, mortality rates were significantly elevated also in the CRC-MS (11.7%) and CRC-AS (18.9%) groups compared to controls (0.0%). Conversely, non-C-carriers showed negligible mortality rates across all groups.

In light of the increased risk of venous thromboembolism (VTE), including Pulmonary Embolism (PE) and Deep Vein Thrombosis (DVT), among CRC patients, monitoring for thromboembolic events is principal. Given the multifactorial nature of VTE, including genetic influences, we examined the association between CDKN2B-AS1 polymorphism, specifically the C-allele, and VTE susceptibility in CRC patients (Table IIB2). Significantly elevated VTE rates were observed in CRC patients with mid-stage and advanced stage disease compared to the control group. Moreover, C-carriers exhibited significantly heightened VTE rates in both CRC-MS and CRC-AS groups compared to controls. Conversely, noncarriers showed lower VTE rates across all groups. Elevated levels of D-dimer and fibrinogen, along with shortened aPTT, were notably higher in CRC-AS patients compared to the control group, supporting heightened coagulation activity.

When comparing CRC staging to the control for overall survival (OS) and the incidence of VTE, Kaplan Meier curves for survival revealed significant differences associated with C-carriers, rather than non-C carriers (Table III-A). Kaplan–Meier analyses demonstrated notable decreases in OS and a higher incidence of VTE, particularly evident in CRC-AS. Across the total population, there were significant decreases in OS (Log-rank=61.76, P<.0001), with more pronounced effects observed in patients who developed VTE (Log-rank=18.35, P<.0001), accompanied by a notably higher incidence of VTE (Log-rank=61.29, P<.0001). Among C-carriers, Kaplan–Meier analysis showed significant decreases in OS for the entire population (Log-rank=28.79, P<.0001) and patients who developed VTE (Log-rank=12.99, P<.005), coupled with an elevated incidence of VTE (Log-rank=19.63, P<.001).

**Table III:**
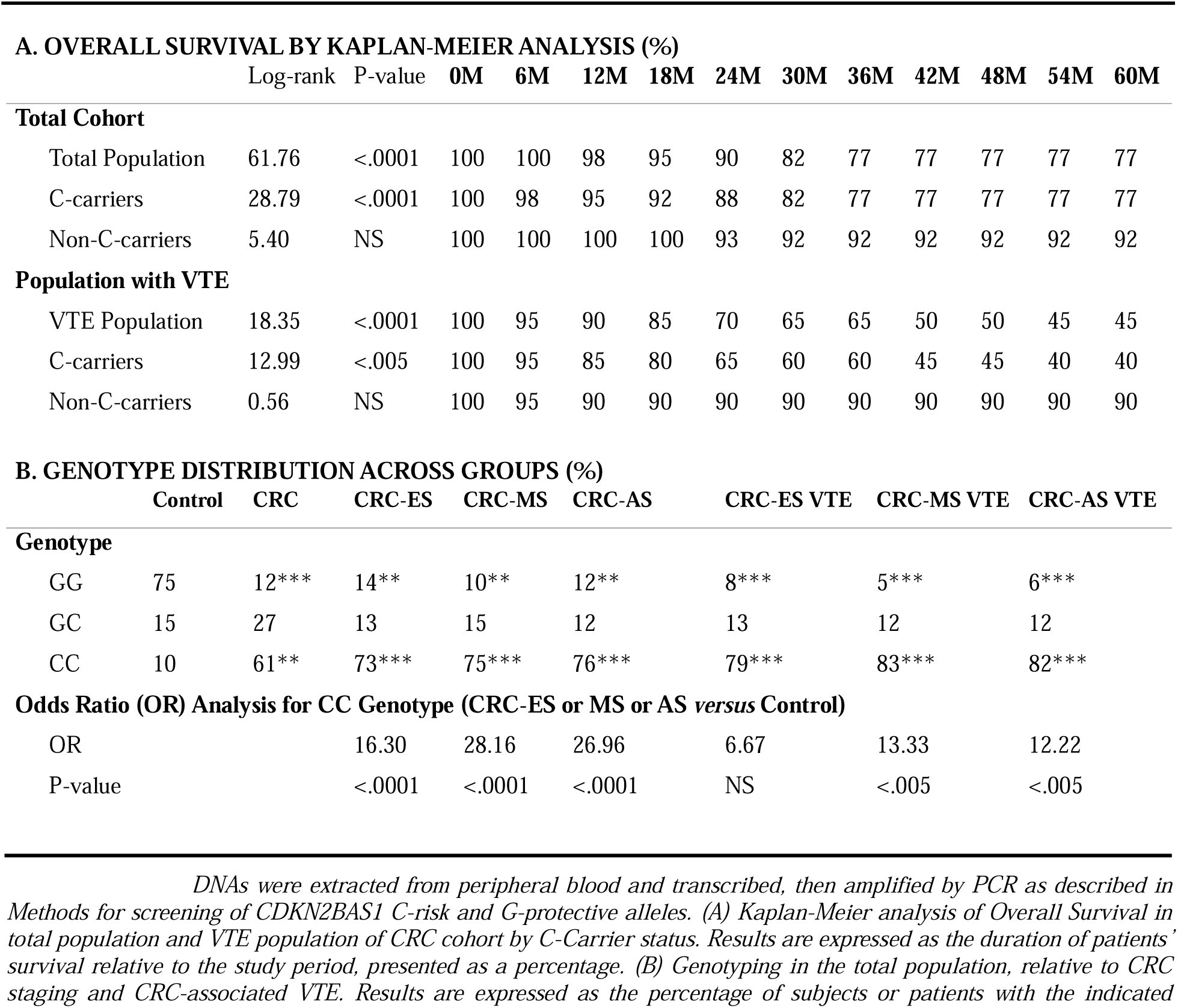
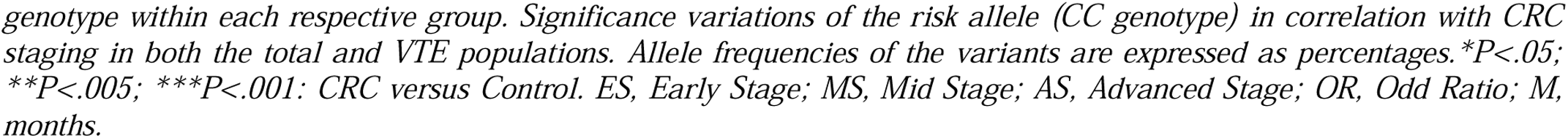
CDKN2B-AS1 Genotyping and Kaplan-Meier Analysis in CRC Patients.

Regarding the median overall survival time (mOS), a 60-month analysis revealed that for CRC patients, mOS ranged from 44 to 54 months, while for those with VTE, it varied from 36 to 49 months—contrasting with control values (54.5-60 and 51.5-53.5 months, respectively). Remarkably, the most pronounced differences in mOS emerged within CRC-AS C-carriers, dropping to 44 months—markedly lower than the 54 months noted in non-C-carriers. These differences were further accentuated among CRC-VTE patients, where mOS declined to 36 months in C-carriers, contrasting to 49 months in non-C-carriers.

These findings highlight the pivotal role of C-carrier status in shaping survival outcomes and increased risk of VTE. Understanding these relationships may contribute to improved risk assessment and management strategies for VTE in CRC patients.

### 3.3. CDKN2B-AS1 Genotyping in CRC

CRC patients predominantly exhibit C-carrier status with homozygous genotype, irrespective of CRC stage, and underscore a strong association with VTE (Table III-B). In contrast to controls, where GG genotype is predominant, CRC patients revealed a predominance of CC genotype (60.7% prevalence). Remarkably, the CC genotype was significantly prevalent across all stages of CRC, showing consistent frequencies in CRC-ES, CRC-MS, and CRC-AS [73.4%, 75.3%, and 75.7% respectively]. Furthermore, among CRC patients who developed VTE (CRC-VTE), the incidence of the C-allele rose to higher levels, with a notable predominance of the CC genotype in CRC-ES, CRC-MS, and CRC-AS [78.6%, 83.3%, and 81.8%, respectively], without significant variations among CRC stages.

### 3.4. Circulating Immune Cells (CICs) Profile in CRC C-carriers

The investigation of Circulating Immune Cells (CICs) in CRC patients carrying the CDKN2B-AS1-Callele underscores their pivotal role in immune dysregulation and inflammation during CRC carcinogenesis. Understanding their dynamics in relation to C-allele offers valuable insights into potential biomarkers and pathways influencing CRC development and prognosis.

Initially, a comprehensive assessment including Complete Blood Count (CBC), differential, and Peripheral Blood Mononuclear Cells (PBMCs) was conducted (Table IV-A). Significant variations were correlating with CRC staging, with the most pronounced changes seen in the CRC-AS group. Within CRC-AS, notable increases were noted in total PBMCs, monocytes, and neutrophils [1.95, 2.94, and 2.89-fold increase respectively], alongside a substantial decrease in lymphocyte count (−57.0%). The Neutrophil-to-Lymphocyte Ratio (NLR), indicative of inflammation, increased significantly (4.95-fold) compared to the control. Notably, different patterns may be exhibited within lymphocyte subsets, including cytotoxic, regulatory, and helper cells, warranting careful consideration.

**Table IV:**
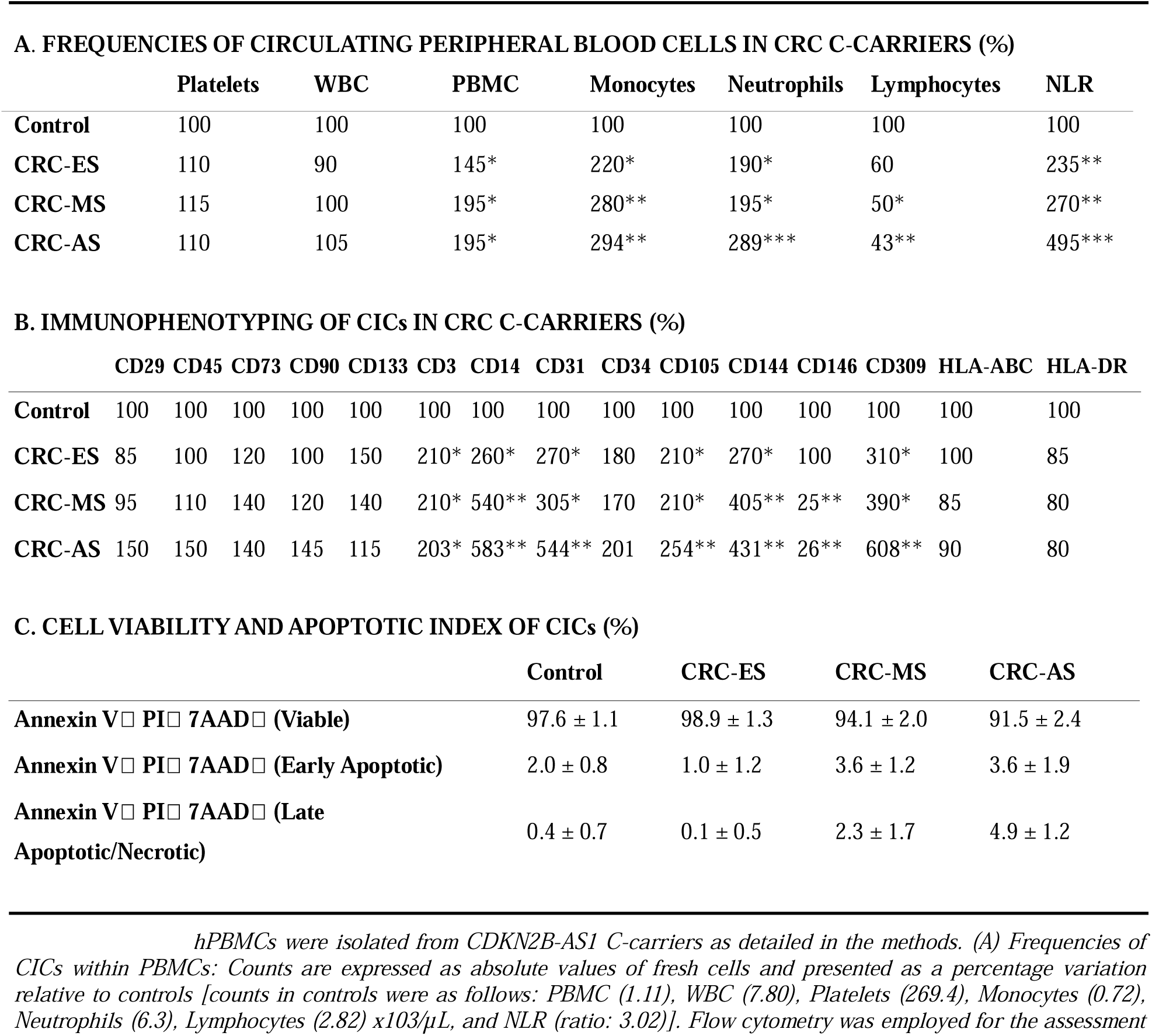
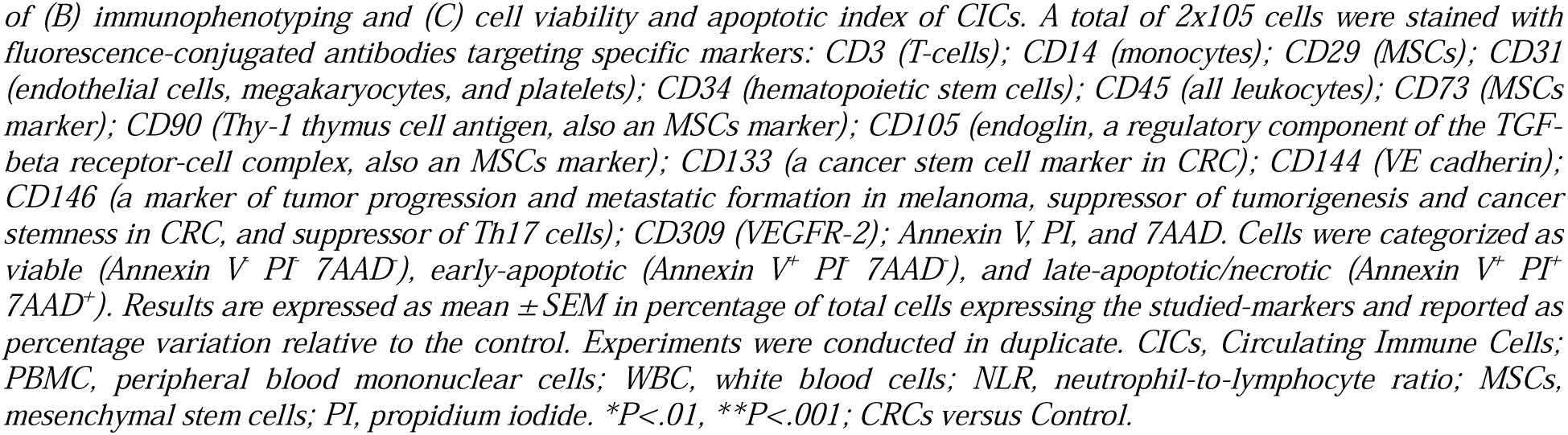
Frequencies and Immunophenotyping of CICs in CDKN2B-AS1 C-carriers with CRC.

Next, circulating PBMCs underwent immunophenotyping (Table IV-B). Compared to the control, significant increases were observed in cells expressing CD3^+^ (2.03-fold), CD14^+^ (5.83-fold), CD31^+^ (5.44-fold), CD34^+^ (2.01-fold), CD105^+^ (2.54-fold), CD144^+^ (4.31-fold), and VEGFR2^+^ (6.08-fold) within the CRC groups, with CRC-AS exhibiting the highest counts. Conversely, a significant decrease of −74.4% was noted in CD146^+^ cells. No significant variations were observed in the counts of CD29^+^, CD45^+^, CD73^+^, CD90^+^, and CD133^+^ cells. Additionally, although decreases in the number of cells expressing HLAABC/DR (MHC) antigens were observed in CRC groups, these decreases did not reach statistical significance. Viability and apoptotic index, assessed using Annexin V/PI/7AAD, showed no significant changes with CRC staging (Table IV-C). The data revealed predominant increases in the abundance of two hematopoietic progenitor cells, CD14^+^ monocytes, and VEGFR-2^+^, primarily involved in angiogenesis, in advanced CRC stages. Elevated CD144^+^ expression suggests potential involvement in cancer progression, indicating acquisition of angiogenic potential by PBMCs. Significant increases were also observed in CD31^+^ cells, which are known for their role in platelet activation and may increase thrombotic risk through the formation of monocyte-platelet aggregates. Conversely, the decrease in CD146^+^ cells, known for their tumor-suppressive roles (in suppressing tumorigenesis, cancer stemness, and Th17 cells), may promote tumor progression as suppression diminishes with CRC progress. These findings collectively support the hypothesis that these alterations may contribute to cancer cells evading the immune system and enhancing their metastatic potential.

Different subclasses of circulating CD3^+^ T-cells, including CD3^+^CD4^+^ (helper T-cells), CD3^+^CD8^+^(cytotoxic T-cells), CD3^+^CD25^+^ (regulatory T-cells), and CD3^+^CD38^+^ (activated T-cell subset), as well as CD3^-^CD56^+^ (NK cells) were investigated, to explore their potential prognostic significance in CRC, specifically in relation to the CDKN2B-AS1 C allele (Table V).

**Table V:**
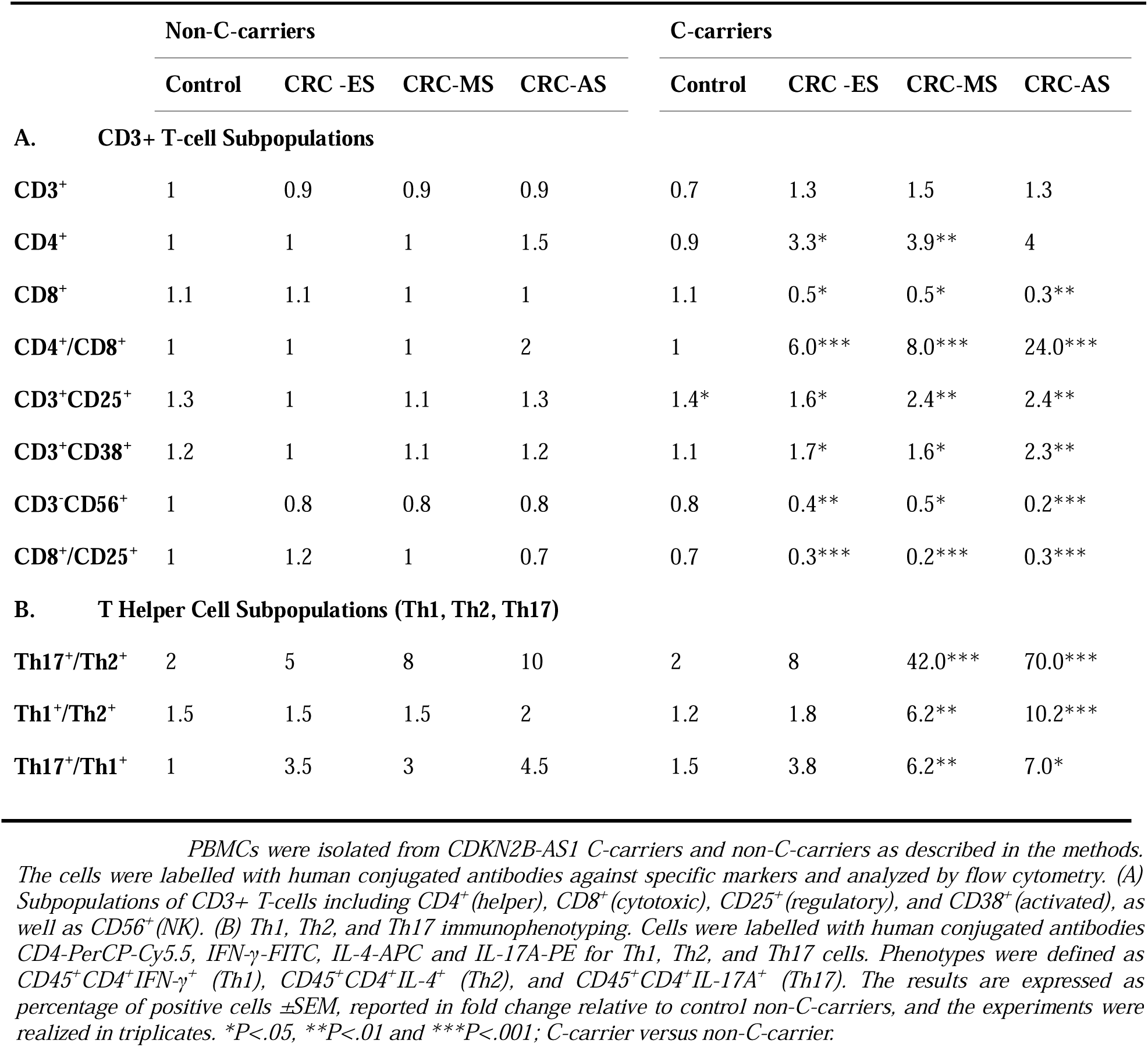
Immunophenotyping of Circulating T-cells in CRC Patients Carrying the CDKN2B-AS1 Risk Allele.

Marked variations were evident in C-carrier CRC-AS patients; notably, significant elevations were observed in CD4^+^, CD25^+^, and CD38^+^ (6.45, 2.44, and 2.36-fold, respectively) relative to controls. Conversely, significant reductions were noted in CD8^+^ T-cells (−75.4%) and CD3^-^CD56^+^ NK cells (70.0%). The CD4^+^/CD8^+^ ratio significantly increased to a 24.2-fold, while the CD8^+^/CD25^+^ ratio, recognized as a predictive marker for survival in CRC patients, decreased significantly (−86.1%) (Table VA). An increased CD4^+^/CD8^+^ ratio could suggest a chronic inflammation or immune dysregulation, potentially promoting tumor growth and progression, while a decreased CD8^+^/CD25^+^ ratio may indicate immunosuppression and immune evasion mechanisms favoring tumor progression. The overall impact of these systemic immune alterations on tumor progression depends on various factors, including the tumor microenvironment and the balance of pro-tumor and anti-tumor immune responses. Thus, further investigation is necessary to understand the precise implications for CRC progression and prognosis.

Notably, significant variances in T-helper cells, crucial components of the immune contexture linked to CRC prognosis in human CRC [38], were observed among circulating T-cells. Given their relevance to inflammation, strongly associated with CRC [39], we explored the potential link between these T-helper cells and CRC, particularly concerning the CDKN2B-AS1 risk C-allele. Inflammatory phenotypes (Th1/Th2/Th17) were evaluated, where Th1 and Th17 cells are proinflammatory, while Th2 cells are anti-inflammatory [40]. Our results (Table VB) demonstrated a significant increase in Th1 and Th17 cells accompanied by a marked decrease in Th2 cells, particularly among CRC-AS C-allele carriers. The Th17^+^/Th1^+^ ratio increased significantly by 7.0-fold, and the Th17^+^/Th2^+^ ratio even more pronounced by 71.8-fold. Additionally, the Th1^+^/Th2^+^ ratio increased significantly by 10.3-fold. These findings underscore the critical role of Th-cells and provide evidence of an inflammatory status linked to the presence of the risk C-allele, mediated by Th1 and Th17 cells.

### 3.5. Inflammatory Biomarkers and Cytokine Levels in CRC C-carriers

Inflammation dysregulation impacts CRC-related biomarkers, including among others elevated serum LDH levels and the production of circulating cytokines. These factors are associated with poor clinical outcomes and therapy resistance, emphasizing the need for their assessment in CRC patients stratified by CDKN2B-AS1 C-carrier status (Table VI).

**Table VI:**
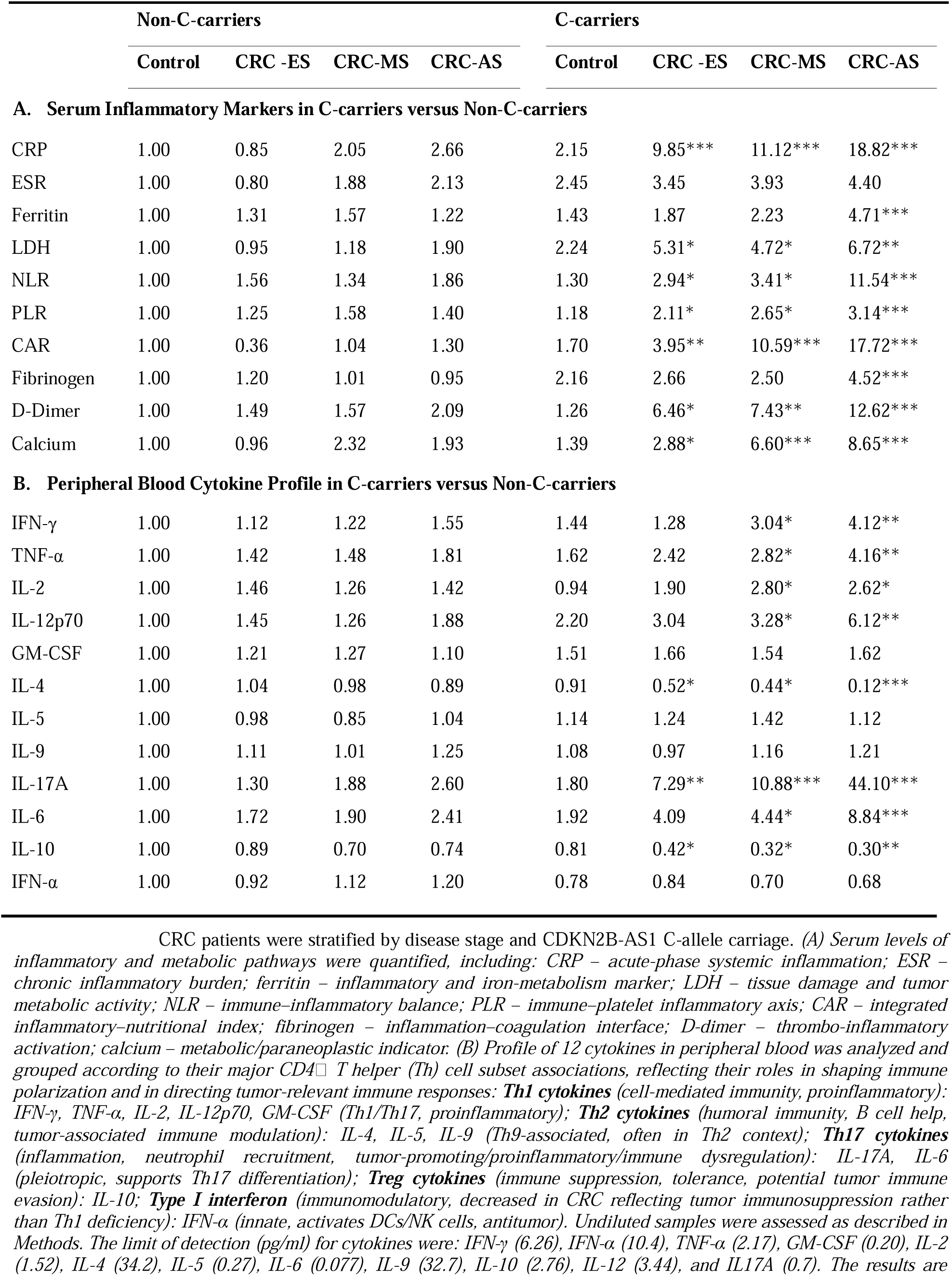
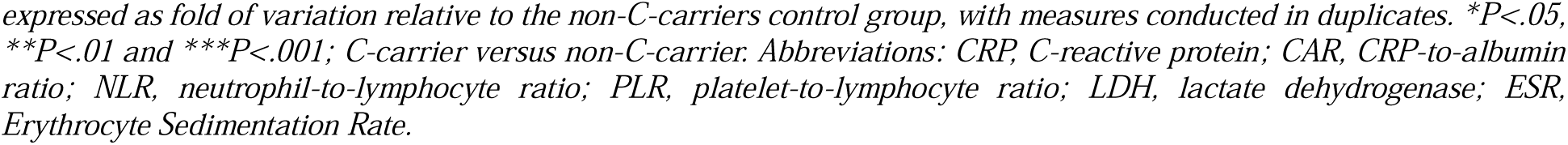
Inflammatory Markers and Cytokines Levels in CRC Patients Carrying the CDKN2B-AS1 Risk Allele.

Our investigation revealed significant increases in circulating inflammatory markers and inflammation associated markers, such as CRP, CAR (CRP-to-Albumin ratio), NLR (Neutrophil-to-Lymphocyte ratio), PLR (Platelet-to-Lymphocyte ratio), LDH, ferritin, and fibrinogen (Table VI-A). Interestingly, these elevations were more pronounced in advancing stages of C-carrier CRC patients, with fold changes of 18.8, 17.7, 11.6, 3.14, 6.79, 4.7, and 4.5, respectively. Additionally, we explored D-dimer levels, a hypercoagulability associated inflammation marker known for its role in cancers. In CRC-AS C-carriers, D-dimer levels significantly increased by 12.6-fold. Furthermore, circulating calcium levels (also known as Factor IV), previously linked to CRC prognosis [41] and correlated with lower survival rates, have been studied. Notably, our data revealed significantly elevated circulating calcium levels in C-carrier CRC patients: CRC-ES (2.9-fold), CRC-MS (6.6), and CRC-AS (8.7-fold increases).

The analysis of circulating cytokine profiles (Table VI-B) revealed that the levels of pro-inflammatory cytokines (IL-17A > IL-6 > IL12p70 > TNF-α and IFN-γ > IL-2) were significantly elevated in CRC Ccarriers compared to non-C-carriers. These levels increased notably in advancing stages, reaching fold changes of 44.1, 8.84, 6.12, 4.16, 4.12, and 2.62, respectively. Conversely, anti-inflammatory cytokines IL-4 and IL-10 exhibited significant decreases (−86.9% and −63.0%, respectively), which may impede tissue repair and homeostasis, potentially contributing to inflammation-induced tumorigenesis. No significant variations were observed for INF-α, GM-CSF, IL-5, and IL-9. The dysregulated cytokine balance in CRC could play a crucial role in the inflammatory microenvironment associated with disease progression, with potential implications for targeted therapeutic approaches.

### 3.6. Circulating Endothelial Progenitor Cells (EPCs) and Angiogenesis in CRC C-carriers

EPCs, integral to neovascularization, have drawn interest in cancer biology, particularly their potential link to CRC. Their involvement in tumor angiogenesis suggests a role in tumor progression and metastasis, making them prospective biomarkers for disease detection and staging [42]. Neoplastic site vascularization is crucial for cancer survival and progression, with EPCs proliferating and differentiating into active endothelial cells, influencing vascular homeostasis and vasculogenesis. Notably, circulating EPC levels are elevated in CRC patients [43]. Therefore, we aimed to determine whether any variations occur in CRC patients, predisposed or not to the CDKN2-AS1 C-allele.

Distinctive phenotypes of EPC subpopulations were studied based on cells having features—circulating or tissue-adherent, including interactions with tumor sites. Given our focus on circulating cells, explored features critical for cell migration and recruitment to tissue injury sites, especially the tumor microenvironment. Based on specific markers (CD31, CD34, CD45, CD144, KDR (VEGFR2), and CXCR4), we identified two EPCs groups: Circulating Non-Tissue-Adherent EPCs (NTA-EPCs) and Tissue-Adherent EPCs (TA-EPCs). Also, we distinguished two subpopulations in TA-EPCs based on the expression of the CD146, an endothelial cell marker suppressor of tumorigenesis. In summary, our study identified three distinct EPC phenotypes within the CRC patient population (Table VII).

**Table VII:** Endothelial Progenitor Cells (EPCs) Phenotypes in CRC Patients.

| EPCs PHENOTYPES | Marker Expression <sup>(1)</sup> |
| --- | --- |
| **Circulating Non-Tissue-Adherent EPCs (NTA-EPCs)** |  |
| Phenotype "A" | CXCR4<sup>+</sup>/KDR<sup>+</sup>/CD31<sup>+</sup>/CD34<sup>+</sup>/CD45<sup>-</sup>/CD144<sup>-</sup>/CD146<sup>-</sup> |
| **Circulating Tissue-Adherent EPCs (TA-EPCs)** |  |
| Phenotype "B", TA-EPCs lacking CD146 | CXCR4<sup>+</sup>/KDR<sup>+</sup>/CD31<sup>+</sup>/CD34<sup>+</sup>/CD144<sup>+</sup>/CD45<sup>-</sup>/CD146<sup>-</sup> |
| Phenotype "C", TA-EPCs expressing CD146 | CXCR4<sup>+</sup>/KDR<sup>+</sup>/CD31<sup>+</sup>/CD34<sup>+</sup>/CD144<sup>+</sup>/CD146<sup>+</sup>/CD45<sup>-</sup> |

As shown in Table VIII, CRC patients with the C-carrier genotype exhibited significantly elevated levels of circulating EPCs expressing both phenotype A and B. Specifically, a 3.73-fold increase for phenotype A was observed in CRC-AS, while phenotype B holds a substantial 7.1-fold increase in CRC-AS compared to the control group. Conversely, a marked reduction in the levels of circulating EPCs expressing phenotype C, characterized by TA-EPCs expressing CD146, reached highest decreases in CRC-AS (80.8%,P<0.001).

**Table VIII:** EPC Surface Marker Analysis in Colorectal Cancer: Influence of CDKN2B-AS1 C-Carrier Status on EPC Phenotypic Distribution.

| EPC Phenotype | CRC Group | non-C-carriers | C-carriers | Significance |
| --- | --- | --- | --- | --- |
| **Phenotype A (NTA-EPCs)** | Control | 1.00 | 1.46 | NS |
|  | CRC-ES | 1.14 | 2.09 | NS |
|  | CRC-MS | 2.27 | 3.61 | NS |
|  | CRC-AS | 1.02 | 5.46 | * |
| **Phenotype B (TA-EPCs)** | Control | 1.00 | 1.90 | NS |
|  | CRC-ES | 0.91 | 4.20 | ** |
|  | CRC-MS | 1.76 | 5.78 | ** |
|  | CRC-AS | 1.71 | 13.51 | *** |
| **Phenotype C (TA-EPCs)** | Control | 1.00 | 0.72 | NS |
|  | CRC-ES | 0.82 | 0.67 | NS |
|  | CRC-MS | 0.92 | 0.33 | ** |
|  | CRC-AS | 0.76 | 0.14 | *** |

### 3.7. Targeted Therapy in Focus: Palbociclib, a CDK4/6 Inhibitor

While chemotherapy remains a cornerstone of CRC treatment, its efficacy in individuals with the C-carrier aggressive phenotype may be suboptimal compared to those without the C-allele. To address this challenge, we adopted a combinatorial approach involving targeted therapy and chemotherapy, focusing on promising agents like Palbociclib (PAL), a CDK4/6 inhibitor known to target the CDK4/6 signaling pathways regulated by the lncRNA CDKN2B-AS1. In our study, CRC-AS patients received chemotherapy CAPOX, which combines capecitabine and oxaliplatin, either with or without Palbociclib. In our cohort of 74 CRCAS patients, 52 patients initially received chemotherapy CAPOX. However, some individuals left the study due to various reasons such as loss of contact, opting for treatment at other hospitals, declining second-line therapy, or leaving the country for socio-economic reasons. Additionally, post-baseline imaging was not conducted for 6 patients, rendering them not evaluable for analysis. As a result, our final analysis included 46 patients who received CAPOX, with 34 not receiving Palbociclib (PAL) treatment and 12 receiving PAL as second-line therapy. Adverse events (AEs) related to treatments were assessed, with additional, non-significant occurrences observed in patients who received CAPOX+PAL compared to those who received CAPOX alone. These included residual side effects, hypertension, headache, dry skin, back pain, and diarrhea.

The evaluation of treatment efficacy included the primary clinical endpoint of Progression-Free Survival (PFS), as well as secondary endpoints such as tumor regression and the levels of key CRC-related biomarkers reflecting tumor burden, proliferation, angiogenesis, and immune status, including CEA, CA 19-9, EGFR, VEGF-A, PD-1, CTLA-4, and Ki-67 (Table IX).

**Table IX:** Effect of Palbociclib Treatment on Progression-Free Survival and Biomarker Response in CRC-AS Patients Carrying the C-Allele.

|  | n | mPFS | Log-rank | P-value |
| --- | --- | --- | --- | --- |
| PAL | 12 | 22 months | 6.18 | 0.0129 |
| Baseline Control | 34 | 9 months |  |  |

|  | CEA | CA 19-9 | EGFR | VEGF-A | PD-1 | CTLA-4 | Ki-67 |
| --- | --- | --- | --- | --- | --- | --- | --- |
| non-C-carriers | -53.2** | -43.5** | -10.1 | -39.8* | -30.4* | -21.4* | -73.4*** |
| C-carriers | -15.1 | -10.1 | -4.8 | -8.9 | -7.8 | -12.0 | -24.2 |

First, when comparing PAL to the control treatment for the PFS, Kaplan-Meier curves exhibited significant differences associated with treatment (Log-rank=6.18, P=0.0012) (Table IXA), indicating improvement. In fact, the median of PFS significantly increased from 9 months in control group to 20 months in PAL group.

The response to PAL treatment differed between C-allele carriers and non-carriers, with important implications for therapeutic outcomes. At 1-year post-treatment, CRC-AS patients in both groups showed reductions in key tumor and immune biomarkers (CEA, CA 19-9, EGFR, VEGF-A, PD-1, CTLA-4, and Ki-67). Importantly, the magnitude of biomarker decrease was generally greater in non-C-carriers, ranging from approximately 10–75%, compared to 5–24% in C-carriers, indicating a stronger biomarker response to PAL in non-C-carriers, representing a high-responder group, whereas C-carriers showed attenuated biomarker reductions, corresponding to a low-responder or PAL-resistant group (Table IXB).

Overall, these findings shed light on the varying effects of PAL depending on the LncRNA genetic predisposition of CDKN2B-AS1, especially on tumor markers and immune checkpoint markers, coupled with a robust significant reduction in the cell proliferation marker Ki-67. These insights are pivotal in tailoring treatment strategies, suggesting the need for personalized approaches based on genetic profiles to optimize therapeutic outcomes and improve patient prognosis.

## 4. Discussion

This study identifies the LncRNA CDKN2B-AS1 C-variant as a “pan-cancer risk biomarker with prevalent occurrence”, revealing its ubiquitous presence across various cancer types and its role as a significant risk indicator, particularly solid carcinomas. Its highest occurrence is noted in gastrointestinal cancers, followed by lung, genitourinary, breast cancer, and others. Moreover, it enhances susceptibility to colorectal cancer (CRC), with the most significant effects observed in patients at advanced stages (CRC-AS). A notable decrease in overall survival and an increased incidence of thromboembolic events were observed, concomitantly with higher frequencies of thromboembolic-associated polymorphisms such as FGB (A), FV (Leiden), and FII (A), and decreased frequency of the protective FXIII (V34L). CRC-AS C-carriers exhibited an immunosuppressive phenotype characterized by a proinflammatory state and impaired immunosurveillance in both circulating immune cells (CICs) and tumor-infiltrating immune cells (TIICs), alongside an increase in angiogenic endothelial progenitor cells (EPCs), potentially contributing to CRC progression and angiogenesis. Significant increases were noted in circulating peripheral blood cells, including PBMCs, monocytes, and neutrophils, accompanied by a decrease in lymphocyte levels. Among PBMCs, there was a notable rise in VEGFR-2, CD14^+^, CD31^+^, CD144^+^, CD105^+^, and CD34^+^, while tumor suppressor-CD146^+^ cells showed a significant decrease. Shifts in T-cell populations were observed in the CICs, with increases in CD4^+^, CD25^+^, and CD38^+^ subsets, along with decreases in circulating CD8^+^ T-cells and CD3^-^CD56^+^ NK cells. Notably, there were increases in Th1 and Th17 subsets, while Th2 saw a drastic decrease. Elevated levels of inflammatory and coagulation markers, as well as proinflammatory cytokines, were observed, contrasting with decreased anti-inflammatory cytokines. Interestingly, circulating EPCs were markedly affected, with levels of non-tissue-adherent EPCs (NTA-EPCs) and tissue-adherent EPCs

(TA-EPCs) lacking CD146 found to be higher, while TA-EPCs expressing CD146 were attenuated. Furthermore, TIICs showed a shift in T-regulatory cells and T-helper cells, with a reduction observed in dendritic, cytotoxic, and NK cells. Additionally, the expression of key CRC-associated biomarkers, including PD-1, CTLA-4, CEA, CA 19-9, EGFR, VEGF-A, and Ki-67, exhibited an increase. A higher prevalence of BRAF mutations and microsatellite stability (MSS) was observed in CRC-AS C-carriers, with no detected KRAS mutations, indicating an aggressive phenotype. Targeted therapy utilizing the

CDK4/6 inhibitor Palbociclib (PAL) demonstrated a more favorable response in CRC-AS non-C-carriers compared to C-carriers, as evidenced by tumor regression and changes in cancer, angiogenic, and immune checkpoint biomarkers, highlighting the aggressive nature of the CRC C-carrier phenotype. Notably, progression-free survival (PFS) increased from 9 to 20 months following PAL treatment at the 1-year outcome.

Globally, the occurrence of the CDKN2B-AS1 C-variant varies among populations, typically around 0.42. Our findings are consistent with the published data, as we observed a frequency of 0.55 in the total population of 882 screened subjects, including both cancer-free individuals and those with cancer.

In addition, ANRIL rs1333049 (G > C) and miR-146A rs2910164 (C > G) have been reported as potential predictive biomarkers for improved prognosis in lung cancer patients receiving platinum-based therapy [44]. Additionally, ANRIL elevated levels have been linked to lower survival rate and a higher metastatic rate in CRC patients [10]. Our study provides insights into the prevalence of the C-allele across various cancer types, highlighting its greater involvement in solid tumors compared to hematological malignancies, with the highest levels observed in gastrointestinal cancers, particularly CRC. Seifi et al. reported that carriers of the CDKN2B-AS1 “C” risk allele had a higher risk of developing breast cancer [9,45]. Additionally, ANRIL rs1333049 (C>G) was found to be significantly associated with an increased risk of cancer, suggesting its potential as a new predictive marker for cervical carcinogenesis [46]. rs7017386 (C>G) was previously associated with CRC risk [47]. Furthermore, ANRIL rs1333049 and miR146A rs2910164 (C>G) have been identified as useful indicators for predicting a better prognosis in lung cancer patients undergoing platinum-based treatment [44]. Elevated levels of ANRIL have also been linked to lower survival rates and higher metastatic rates in CRC patients [10]. In fact, elevated levels of ANRIL, have been associated with increased CDK4/6 activity. ANRIL is involved in epigenetic regulation and can influence the expression of genes involved in cell cycle control, including CDK4 and CDK6. Therefore, increased levels of ANRIL can contribute to tumor progression by promoting CDK4/6 activity and enhancing tumor cell proliferation, thus ANRIL stands as a fascinating and multifaceted molecule with diverse implications in several types of cancer [14]. Interestingly, our C-carriers have high levels of CDK4/6 activity.

Increased incidence of VTE was noted in CRC-AS C-carriers. In fact, the role of CDKN2B-AS1 in thromboembolism is well established in a variety of diseases [18,48], most notably vascular diseases. The link between cancer and VTE has been well documented; in fact, cancer patients with abnormal blood clotting mechanisms are at significantly increased risk of thromboembolism, particularly those receiving systemic chemotherapy [49]. Our findings revealed elevated levels of D-dimer, fibrinogen, and shortened aPTT in agreement with increased thromboembolic events, highlights the impact of the C-variant in CRC susceptibility. These thromboembolic dysregulations controlled by cancer cells including among others production of inflammatory and angiogenic cytokines, are confirmed in our cohort study [22,25,49].

ANRIL is thought to be involved in the formation of the tumor microenvironment matrix and may aid in cancer progression, so to be involved in tumor metastasis [9]. The tumor microenvironment is crucial in understanding immunosurveillance and immunoediting in cancer [20,21]. The connection between CDKN2B-AS1 and CICs in CRC showed strong involvement of inflammatory markers. Despite the decrease of total lymphocytes count in CRC groups, significant variations occurred in T-cells subsets. The CD8+ cytotoxic T-cells, key effectors of the cell-mediated immune response, are usually suppressed in cancers to promote tumor growth [50]; indeed, our results showed clearly a decreased CD8+ T-cells count in the C-carriers. CD4+ T-cells subsets in correlation with genetic predisposition showed a pronounced rise in the proinflammatory Th1 and Th17 contrary to a decrease in the anti-inflammatory Th2; high Th17 clusters have been previously linked to poor CRC prognosis [40]. Immune/inflammatory cells infiltrate practically all human solid tumors as they produce different cytokine subsets. Importantly, overproduction of Th17 cytokine product, IL-17A, marks the early stages of CRC and negatively influences the prognosis of CRC patients [51]. Evidences suggest that overproduction of IL-17A is associated with the production of VEGF and poor prognosis in patients with CRC [52]. The detected proinflammatory profile of circulating cytokines (high levels of IL17A, IL12p70, IL-6, IL-2, TNF-α, and IFN-γ concomitantly with low levels of IL-4 and IL10) in CRC-AS C-carriers agree with the globally reported data.

Vascularization of neoplastic sites is a key characteristic that facilitates the survival and progression of cancer. EPCs undergo proliferation and differentiation into active endothelial cells, playing a vital role in vascular homeostasis and vasculogenesis [53]. In the context of cancer, this can have a dual effect. On the positive side, EPCs contribute to angiogenesis for tissue repair [54]. However, it’s important to note that excessive or aberrant angiogenesis driven by EPCs can promote tumor growth and metastasis by enhancing new vessel formation, highlighting a potential negative aspect [43]. Furthermore, our study, consistent with previous findings, demonstrates elevated levels of circulating EPCs in CRC patients [43]. Interestingly, our study showed dynamics between circulating NTA-EPCs and TA-EPCs, especially with the impact in CRC of an aggressive phenotype expressing CD146^+^. In fact, the significant decrease in CD146 expression in PBMCs, particularly in advanced CRC stages, is a noteworthy finding. CD146 has been associated with suppressing tumorigenesis and cancer progression, and its reduction in CRC patients may indeed suggest a loss of this suppression mechanism. Based on the EPCs phenotypes and the known roles of CD146 in tumor suppression and inhibiting cancer stemness, the increase in EPCs lacking CD146 (EPCs-CD146^-^) may suggest a potential tumor-promoting role, where EPCs may lack the tumor-suppressive functions linked to CD146 expression and could potentially contribute to tumor progression or angiogenesis within the CRC microenvironment [55]. On the other hand, the decrease in EPCs expressing CD146 (EPCsCD146^+^) implies also a potential loss of the tumor-suppressive functions associated here with the absence of expression CD146 which could contribute to an environment more conducive to tumor growth and progression [56].

The dynamics of circulating EPCs in CRC-AS-C-carriers’ patients exploring different phenotypes related to tissue-adherence features or not, gave more evidence of the potent role played by the circulatory system. However, no study has yet explored the role of CDKN2B-AS1 and its genetic variants on the dynamics of EPCs. In fact, the increases in cells expressing the CD144 (VE-cadherin), an endothelial-specific adhesion molecule critical for cell-cell adhesion in endothelial junctions, could indicate support for angiogenesis, thereby promoting tumor growth. One of the main inducers of EPCs mobilization is VEGF [57], highly released from CRC tumor tissues to recruit endothelial cells precursors [58]; hence proving a connection between chronic inflammation, CRC and EPCs. Our data showed higher levels of proinflammatory cells in advanced stages of CRC in C-carriers’ patients with concomitant high levels of VEGF. Indeed, one could predict that the production and lifespan of EPCs could also depend on the individual genotype of CDKN2BAS1 gene.

The assessment of PAL, a CDK4/6 inhibitor, as second-line therapy following chemotherapy CAPOX (capecitabine and oxaliplatin) in CRC-AS patients revealed significant differences in PFS between PAL-treated and control groups. Notably, the PAL group exhibited a median increase in PFS from 9 to 20 months, accompanied by significant tumor regression. Moreover, when stratified by C-carrier status, PAL treatment response profiles indicated significant tumor regression and reductions in CRC-associated biomarkers that differed between C-carriers and non-C-carriers, highlighting the influence of genetic predisposition on therapeutic outcomes. Although tumor regression was observed from TRG3 (minimal response) to TRG1 (partial response) in both groups, the impact was more pronounced in non-C-carriers. These findings underscore the necessity for personalized treatment strategies based on genetic profiles to optimize therapeutic outcomes and improve patient prognosis.

As our results indicate, PAL primarily impacts cell proliferation, as evidenced by the notable decrease in Ki-67 levels. Additionally, there are moderate effects on immune checkpoints, as demonstrated by reductions in CTLA-4 and PD-1 levels. However, our observations indicate no significant impact on angiogenesis, as evidenced by the absence of notable changes in VEGF levels. Specifically, compared to baseline, a more pronounced reduction was observed in non-C-carriers compared to C-carriers.

The partial reversibility observed with Palbociclib suggests that individuals with the C-carrier aggressive phenotype may benefit from dual therapies targeting either (1) both the immune checkpoints and the tumor cell division cycle, or (2) both the VEGF axis and the tumor cell division cycle, rather than solely focusing on the latter. Both approaches hypothesize the use of immunotherapy combined with CDK4/6-targeted therapy. These strategies are supported by reported studies demonstrating the efficacy and challenges of targeting immune checkpoints [59] and angiogenesis [60–62] in CRC treatment. Moreover, kinase inhibitors could further enhance treatment outcomes. For example, Encorafenib, a small molecule BRAF inhibitor targeting the MAPK signaling pathway, has been utilized in combination with the anti-EGFR cetuximab to treat CRC, particularly metastatic CRC with BRAF V600E mutation [63]. The interconnectedness of the MAPK pathway and CDKN2B-AS1 [64] with the CDK4/6 pathway suggests that complementary therapy targeting multiple pathways is essential for addressing the aggressive phenotype observed in CRC-AS C-carriers. A comprehensive therapeutic approach for CRC, especially the C-carrier aggressive phenotype, should target not only the proliferative axis but also angiogenic, immune checkpoint, and other implicated signaling pathways in disease progression.

It is well established that PAL inhibits CDK4/6, preventing the formation of cyclin D-CDK4/6 complexes that aid in Rb phosphorylation, thereby inhibiting cell cycle progression. In our model, where individuals carrying the risk C-allele of CDKN2B-AS1 exhibit elevated CDK4/6 activity, treatment with PAL resulted in marked inhibition of Ki-67. This suggests that the increased CDK4/6 activity associated with the C-allele may contribute to enhanced tumor growth and proliferation. In fact, CDKN2B-AS1, an antisense transcript to the CDKN2B gene encoding p15INK4B, a cyclin-dependent kinase inhibitor that negatively regulates CDK4/6 activity, indirectly affects CDK4/6 activity and cell cycle progression by modulating CDKN2B expression. One potential scenario involves the C-allele of CDKN2B-AS1 decreasing p15INK4B expression, leading to elevated CDK4/6 activity, which promotes cell cycle progression and tumor growth. Alternatively, the C-allele of CDKN2B-AS1 may directly enhance CDK4/6 activity through other mechanisms, independent of p15INK4B regulation. Regardless of the scenario, elevated CDK4/6 activity in C-carriers underscores its pivotal role in carcinogenesis, making it a crucial target for therapy. Inhibiting CDK4/6 with Palbociclib reverses this effect, leading to suppression of cell proliferation and tumor regression.

## 5. Conclusion

In summary, this study investigates the impact of CDKN2B-AS1 polymorphism on various cancers, particularly CRC. It underscores the role of CDKN2B-AS1 in counteracting CDKN2B and its association with thromboembolism. Furthermore, it provides compelling evidence of significant alterations in CICs, EPCs, and TIICs subsets in CRC, influenced by the CDKN2B-AS1 polymorphism and microsatellite stability. Moreover, the study suggests a therapeutic approach to enhance CRC treatment by potentially inhibiting CDK4/6 using PAL as a promising target to mitigate immunosuppression and inflammatory reactivity in CRC. The inflammatory axis leading to tumor progression such as the MAPK-NF-kB or PI3kNF-kB pathways could give more comprehensive mechanistic insights of the LncRNA ANRIL and its risk C-allele in the development of cancer. The observed aggressive phenotype among C-carriers warrants further investigation, potentially suggesting the need for associated immunotherapy such as anti-angiogenic (anti-VEGF) treatment to overcome limitations in the therapeutic effectiveness of PAL. Thus, considering a combined therapy approach could yield improved outcomes. Our novel ongoing clinical trial evaluating the application of precision therapy combining PAL with Bevacizumab (an anti-VEGF agent) is currently active (NCT06307249). Additionally, given the absence of a direct RS1333049 inhibitor, exploring new types of therapies targeting the cell cycle in alternative ways is imperative.

## Data Availability

All data produced in the present work are contained in the manuscript

## Acknowledgments

The authors would like to express their sincere gratitude to the medical director at Haykal Hospital, Dr. Lise Abi Rafeh, for her continuous support and guidance throughout the project. The authors also extend their appreciation to the laboratory supervisor, Mrs. Maya Al Eter, for her invaluable assistance and expertise in overseeing the laboratory processes. Additionally, the authors would like to acknowledge the contributions of the lab technicians, Ms. Alaa Hamwi, the molecular biology technician, and Ms. Jenny Elia, the special chemistry technician, for their meticulous work and technical expertise that were instrumental to the success of this study.

Disclosures:

## Ethics Statement

All clinical investigations involving human samples were carried out in adherence to the principles outlined in the Declaration of Helsinki, as revised in 2008 (http://www.wma.net/e/policy/b3.htm), and it was registered in ClinicalTrials.gov (Identifier: NCT06065592). Written informed consent was obtained from all donors, and sample collection was conducted in accordance with ethical guidelines. The study protocol received approval from the institutional review committee of Haykel Hospital (LUHH21/22/P04) renewed for 2023 (LUHH23/14/P06) and the scientific committee of the Lebanese University (1821/36/M, and 6341/4). These ethical approvals ensure that the study is conducted in a manner that respects the rights, welfare, and confidentiality of the participants. The internship to conduct this thesis in the field of molecular diagnostics has received the approval of Haykel Hospital for the period between February and September 2023, with the reference code ILUL220523_EMS which confirms the hospital’s support and endorsement of the internship and thesis during the specified timeframe, specifically in the area of molecular genetics and Immunology. Animal studies: N/A.

## Author Contributions

Elisa Makdessi (EM): principal investigator, study concept and design, experimental assessment, data collection and analyses, writing and critical revision of the manuscript.

Samar El-Hamoui (SEH): co-principal investigator, molecular genetics assessment, data collecting, technical measurement, analysis and interpretation of data, critical revision of the manuscript.

Fida El-Ayoubi (FEA): co-principal investigator, study concept and design, analysis of data.

Malak Naboulsi (MN): co-principal investigator, medical laboratory analyses, data collection.

David Wehbe (DW): co-principal investigator, hemato-oncologist, patients’ enrollment, therapeutic application.

Noha Ibrahim (NI): co-principal investigator, molecular biology assessment.

Nadine Nasreddine (NN): co-principal investigator, immunobiology assessment.

Nehman Makdissy (NM): Principal Investigator, study concept and design, experimental design, data collection, analyses and interpretation, statistical analysis, writing and critical revision of the manuscript.

## Abbreviations

CDK: cyclin-dependent kinase
CDKN2B: cyclin-dependent kinase inhibitor 2B
CDKN2B-AS1: cyclin-dependent kinase inhibitor 2B anti-sense 1
CICs: circulating immune cells
CIICs: circulating infiltrated immune cells
CRC: colorectal cancer
EPC: endothelial progenitor cells
hPBMCs: human peripheral blood mononuclear cells
LncRNA: long non-coding RNA
MSI: microsatellite instability
MSS: microsatellite stability
NK: natural killer
PCR: polymerase chain reaction
VTE: venous thromboembolism.

